# Mapping the role of tumor necrosis factor-related apoptosis-inducing ligand (TRAIL) and its receptors in chronic kidney disease- a scoping review protocol

**DOI:** 10.1101/2025.08.07.25333262

**Authors:** Hansani Niroshika, Harshi Weerakoon, Nalaka Herath, Kosala Weerakoon, Prasad Jayasooriya

**Author notes:** Corresponding author: (KW). Corresponding author: (HN).

## Abstract

**Background:** Chronic kidney disease (CKD) is increasingly recognized as a global public health concern, impacting about 700 million people worldwide. Due to the presence of both pro- and anti-apoptotic properties, tumor necrosis factor (TNF)-related apoptosis-inducing ligand (TRAIL) can be a double-edged sword in the pathogenesis and progression of CKD. Systematic mapping of the available evidence is pivotal in identifying the exact role of TRAIL in CKD. Therefore, this protocol explains the methodological approach for a scoping review to map evidence regarding the role of TRAIL and its receptors in CKD.

**Methods:** The proposed scoping review will be conducted in line with Arksey and O’Malley’s methodological framework and the Joanna Briggs Institute (JBI) reviewer’s manual. Accordingly, the review will be guided by the five-stage approach, namely (1) identify the research question; (2) identify relevant studies; (3) select studies; (4) chart the data; and (5) collate, summarize, and report the results. Eligibility criteria and search strategy will be formulated based on population, concept, and context (PCC) strategy. Articles published up to July 2025 will be searched using electronic databases, PubMed/MEDLINE, Science Direct, Scopus, CINAHL, and Cochrane Library, while considering the Google Scholar reference lists of review articles as grey literature sources. A formal quality appraisal of the selected studies will be conducted using the mixed-method appraisal tool (MMAT version 2018). The reporting will be done following the Preferred Reporting Items for Systematic Reviews and Meta-Analyses extension for the Scoping Reviews checklist. The literature search is expected to commence after the protocol acceptance process, followed by study selection and data extraction by November 2025. Data synthesis will be completed by February 2026.

**Scoping review registration:** The review protocol is registered at Open Science Framework (OSF) under https://doi.org/10.17605/OSF.IO/AENBX.

## 1. Introduction

Chronic kidney disease (CKD) is defined as kidney damage or glomerular filtration rate (GFR) <60 mL/min/1.73 m2 for 3 months or more, irrespective of the cause (1) is a global health burden affecting more than 700 million people worldwide (2). Hypertension and diabetes mellitus are the most common causes of CKD, while factors like genetics, infections, adverse drug effects, and autoimmunity can also cause the disease (3,4). Regardless of the underlying cause, CKD increases the risk of premature deaths caused by cardiovascular diseases (5). On a histological basis, the disease is characterized by the gradual depletion of different cell types including podocytes, tubular epithelial cells, and endothelial cells resulting in glomerulosclerosis, tubular atrophy, and capillary rarefaction. Simultaneously, maladaptive cells like myofibroblasts and inflammatory cells are recruited and proliferated, causing a significant loss of renal function due to fibrosis (6,7). Tumor necrosis factor (TNF)-related apoptosis-inducing ligand (TRAIL) is a TNF superfamily cytokine (member 10) that is highly likely to be involved in CKD pathology. TRAIL can induce apoptosis selectively in transformed cells with minimal off-target effects (8,9). This selectivity arises from the differential expression of death (TRAIL-R1/DR4 and TRAIL-R2/DR5), decoy (TRAIL-R3/DcR1, TRAIL-R4/DcR2), and soluble osteoprotegerin (OPG) receptors of TRAIL. Death receptors, when activated, trigger apoptotic signaling while decoy and OPG receptors promote cell survival and evade TRAIL-induced apoptosis (10). While its role has been studied extensively as an apoptotic inducer of cancer cells, its application in noncancerous diseases, particularly fibrotic and inflammatory conditions such as CKD, is an emerging area of interest.

In a recent study that highlights its potential role in the pathophysiology of CKD, an inverse relationship between TRAIL levels and mortality risk was observed (11). In patients with CKD, a significant association between low levels of circulating TRAIL and atheromatous plaque progression has been identified, and these results highlighted the possible use of TRAIL as an independent prognostic biomarker to monitor atherosclerosis in patients with CKD (12). Further, some recent experimental animal studies have emphasized the potential role of TRAIL in CKD. For example, an experiment conducted using TRAIL(-/-) ApoE(-/-) and ApoE(-/-) mice fed with a high-fat diet for 20 weeks revealed exacerbated diabetic nephropathy in the presence of TRAIL deficiency (13). On the other hand, TRAIL treatment (twice daily over 12 weeks) seems to remodel the glomerular and tubular morphology, causing improved renal functions in db/db (diabetic) mice (14).

Mechanistically, this ligand appears to influence tubulointerstitial injury, glomerular filtration, and vascular remodeling based on the receptor expression modulated by nature of cytotoxic insult and microenvironment. The balance between proapoptotic and antiapoptotic TRAIL receptors determines whether TRAIL signaling promotes cell death, survival, inflammation, or senescence of renal cells. For example, in early diabetic kidney disease (DKD) TRAIL has been associated with podocyte PANoptosis by expressing higher DR5 levels (15). Concurrently, metabolic stress and advanced glycation end products are shown to promote tubular cell senescence and maladaptive repair by expressing higher DcR2 levels (16,17). Likewise, the precise role of TRAIL and its receptors in CKD of different etiologies remains under investigation and literature remains fragmented with limited synthesis of data.

Therefore, mapping the existing evidence systematically will pave the way to identify the significance of TRAIL in CKD and thereby identify the areas that need to be further explored. For this, we selected scoping review approach since existing studies span across different experimental models, clinical observations, and molecular mechanisms among different CKD subtypes. The outcome review paper aims to summarize and highlight the existing research gaps related to TRAIL/TRAIL-R expression patterns, regulatory mechanisms and associations with clinical outcomes across different CKD etiologies. This scoping review protocol will define the transparent and systematic methodology to identify, chart and synthesize existing evidence.

## 2. Methodology

This review will comply with the methodological framework described by Arksey and O’Malley (18) and revisions made by the Joanna Briggs Institute (JBI) (19). Thus, the review will be developed in five stages (Figure 1).

**Figure 1.**
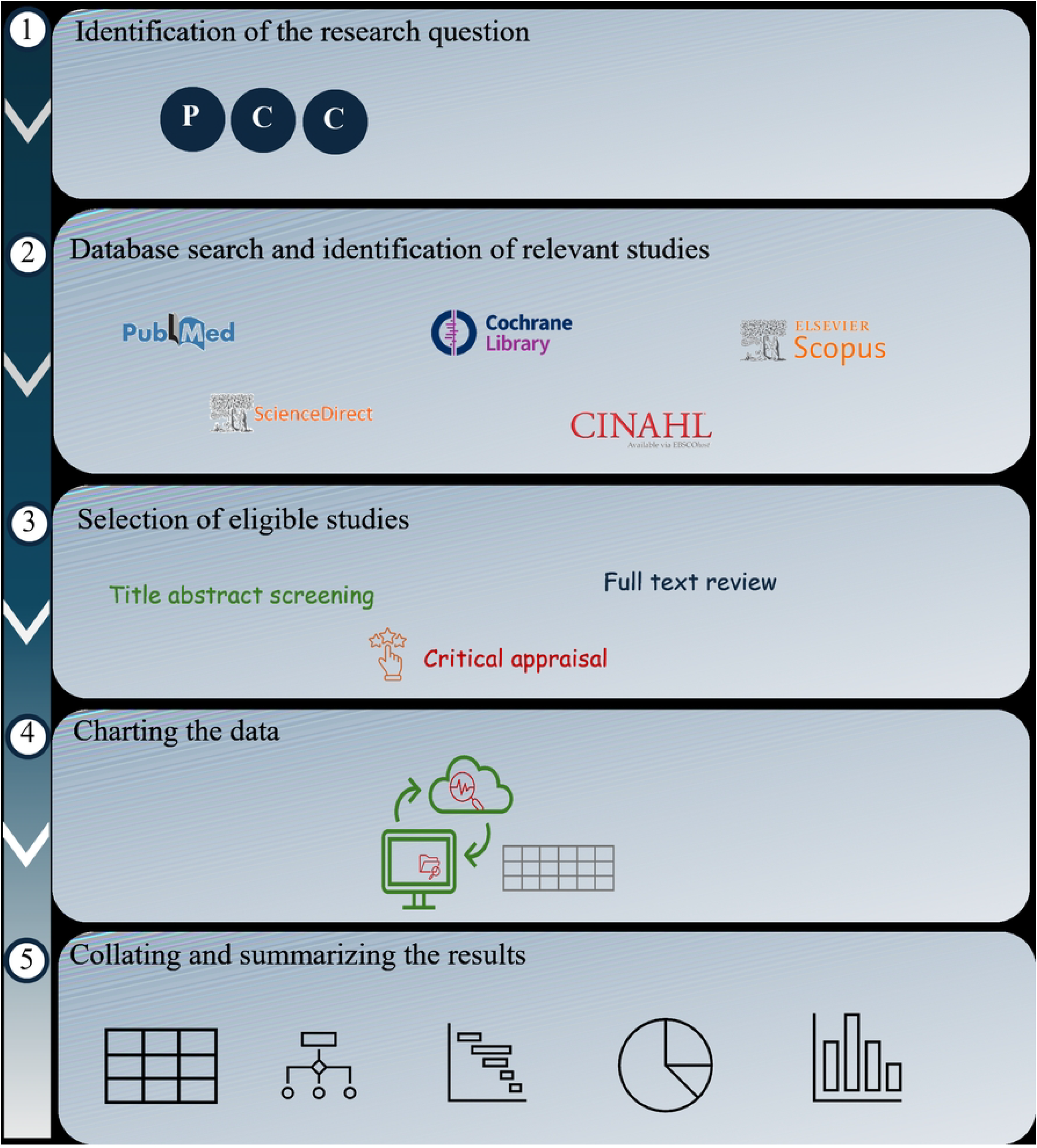
Methodological framework of the scoping review (PCC -population, content, concept)

Scoping review reporting will adhere to the Preferred Reporting Items for Systematic Reviews and Meta-Analyses extension for Scoping Reviews (PRISMA-ScR) checklist (20). The review will be conducted from March 2025 to March 2026, following a structured timeline. The initial phase will involve the development and approval of the review protocol, scheduled for March 2025 to August 2025. The database search is planned to be initiated at the end of August and continue through September 2025. Study selection and data extraction will be completed by November 2025, followed by data synthesis up to February 2026. The manuscript will be drafted and finalized between January and March 2026.

### Stage 1: Identification of research question

The population, concept, and context (PCC) framework (21) is used to formulate the primary review question and to develop the search strategy (Table 1).

**Table 1.** PCC framework used.

| Criteria | Determinants |
| --- | --- |
| **P- Population** | All human studies, animal models, cell lines, and related computational models of CKD |
| **C- Concept** | TRAIL/ TRAIL receptors in CKD research |
| **C- Context** | Not limited to a specific region or country |

The main research question of this review is ‘What are the patterns, mechanisms, and clinical implications of TRAIL and its receptor expression in CKD?’

Areas explored under the above overarching question are,

1. What are the patterns of TRAIL and its receptor expression in CKD?
2. What are the mechanisms underlying the regulation of TRAIL expression in CKD?
3. What are the associations of TRAIL with disease severity, progression, and clinical outcomes in CKD?
4. What are the knowledge gaps in understanding the role of TRAIL across CKDs of different etiologies?

### Stage 2: Database search and identification of relevant studies

The article search will follow the JBI-recommended three-phase search process (21). In phase 1, an initial search will be conducted through PubMed with two main search terms (i) Tumor necrosis factor-related apoptosis-inducing ligand (Tumor necrosis factor-related apoptosis-inducing ligand OR TRAIL OR APO 2L OR APO2L APO-2L OR TNFSF 10 TNFSF10 OR TNFSF-10 OR TNF related apoptosis-inducing ligand OR CD 253 OR CD253 OR CD-253) and (ii) chronic kidney disease (CKD, chronic kidney disease OR CKDu OR chronic renal failure OR CRF OR renal OR kidney OR nephropathy OR glomerulonephritis OR polycystic kidney disease OR chronic interstitial nephritis OR chronic kidney failure OR end-stage renal disease OR ESRD). Then, a second search (phase 2) will be performed with the identified most relevant keywords in phase 1. For this, database-specific search strategies will be implemented on electronic databases, PubMed/MEDLINE, Science Direct, Scopus, CINAHL, and Cochrane Library. All articles published up to July 2025 will be included. In phase 3, reference lists of all identified papers, relevant literature reviews, and the Google Scholar database will be used to find additional and grey literature.

### Stage 3: Selection of eligible studies

Article selection will follow the PCC framework along with the following eligibility criteria to confirm the most relevant studies to review questions.

Inclusion criteria:

1. Quantitative and qualitative human/animal studies or computational models on expression, regulation, mechanisms, and clinical implications of TRAIL in CKD
2. Articles published up to July 2025
3. Reviews in line with the search string will be considered for background details and to trace relevant articles that were not detected in the initial screening process

Exclusion criteria

1. Articles published in languages other than English
2. Articles on renal carcinoma, acute kidney injury, and kidney stones

Depending on the above criteria, a two-step approach will be carried out to filter the articles.

Step 1: Title and abstract screening

The articles retrieved from the database search will be imported into reference management software for deduplication. Two independent reviewers will screen the titles and abstracts and assess whether they meet the inclusion criteria. In case of any disagreement, the opinion of a third reviewer will be obtained.

Step 2: Full text review

Selected studies from Step 1 will be subjected to comprehensive full-text screening to identify their suitability for subsequent data extraction. In selecting and mapping included studies, recommendations from the PRISMA-ScR checklist and PRISMA-P chart will be followed (20). The Rayaan.ai AI-powered systematic review management platform will be used to store and manage data throughout the review process (22).

### Critical appraisal

The mixed-method appraisal tool (MMAT version 2018) will be used to assess the quality of the selected studies. This tool comprises assessment criteria for qualitative, quantitative randomized control trials (RCTs), quantitative non-RCTs, quantitative descriptive, and mixed methods. Each item will be rated on a scale of “yes”, “no”, or “can’t tell” and each study will receive an overall score based on the number of criteria met divided by 5, resulting in scores from 20% (one criterion met) to 100% (all criteria met) (23). The appraisal will be initially conducted by one reviewer and independently assessed by a second reviewer. Any discrepancy will be discussed and resolved by consensus. Scores will be used as a guide to interpret study quality, but no studies will be excluded solely based on low scores.

### Stage 4: Charting the data

Articles selected from the above step will be used for data extraction. Data will be charted using the data extraction framework (Table 2) that captures bibliographic details, study design, population characteristics, TRAIL related biomarkers, and outcomes in CKD progression. This will be conducted under 5 key subject areas, and this preliminary content may be amended as the review progresses.

**Table 2.**
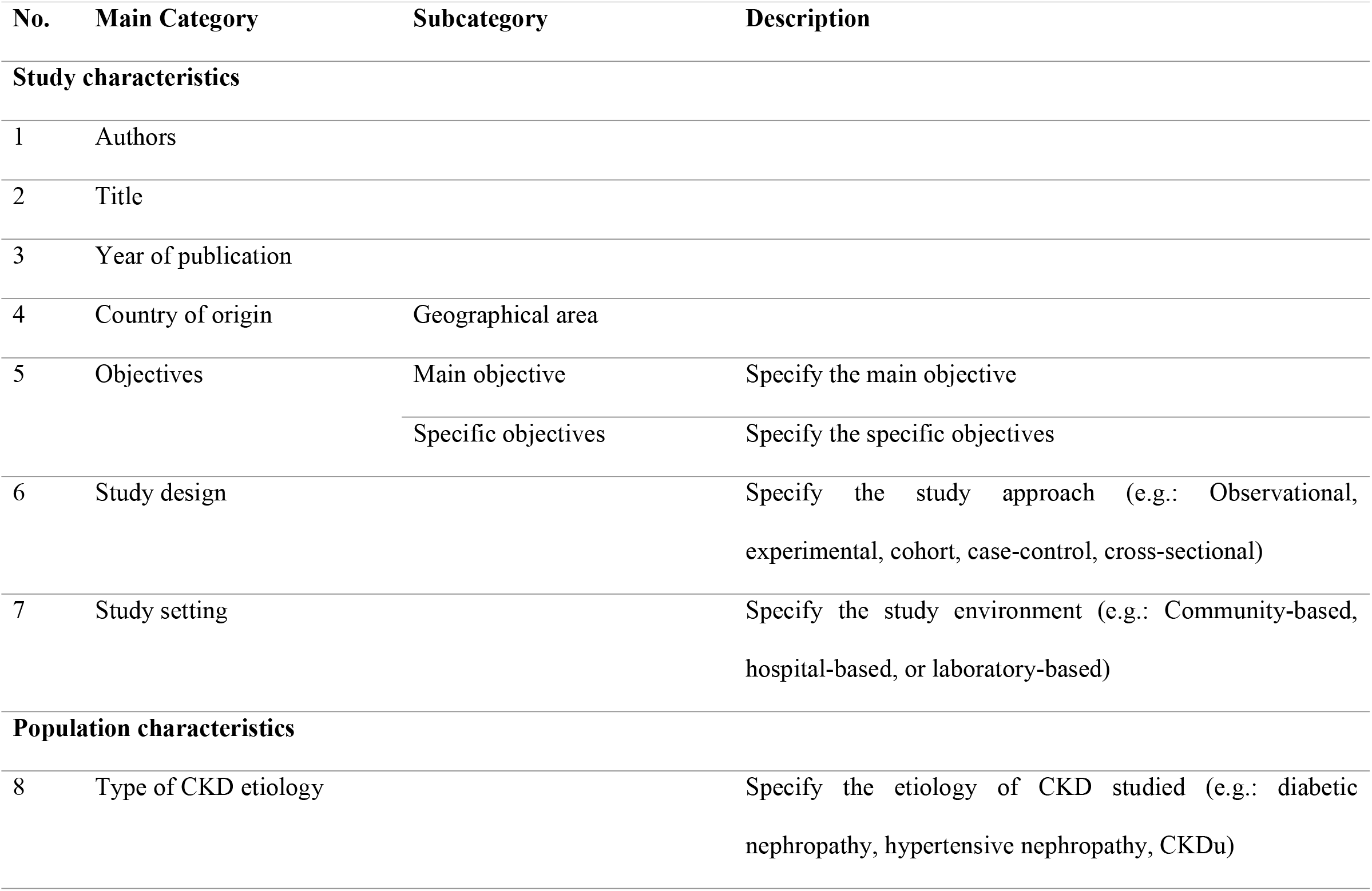
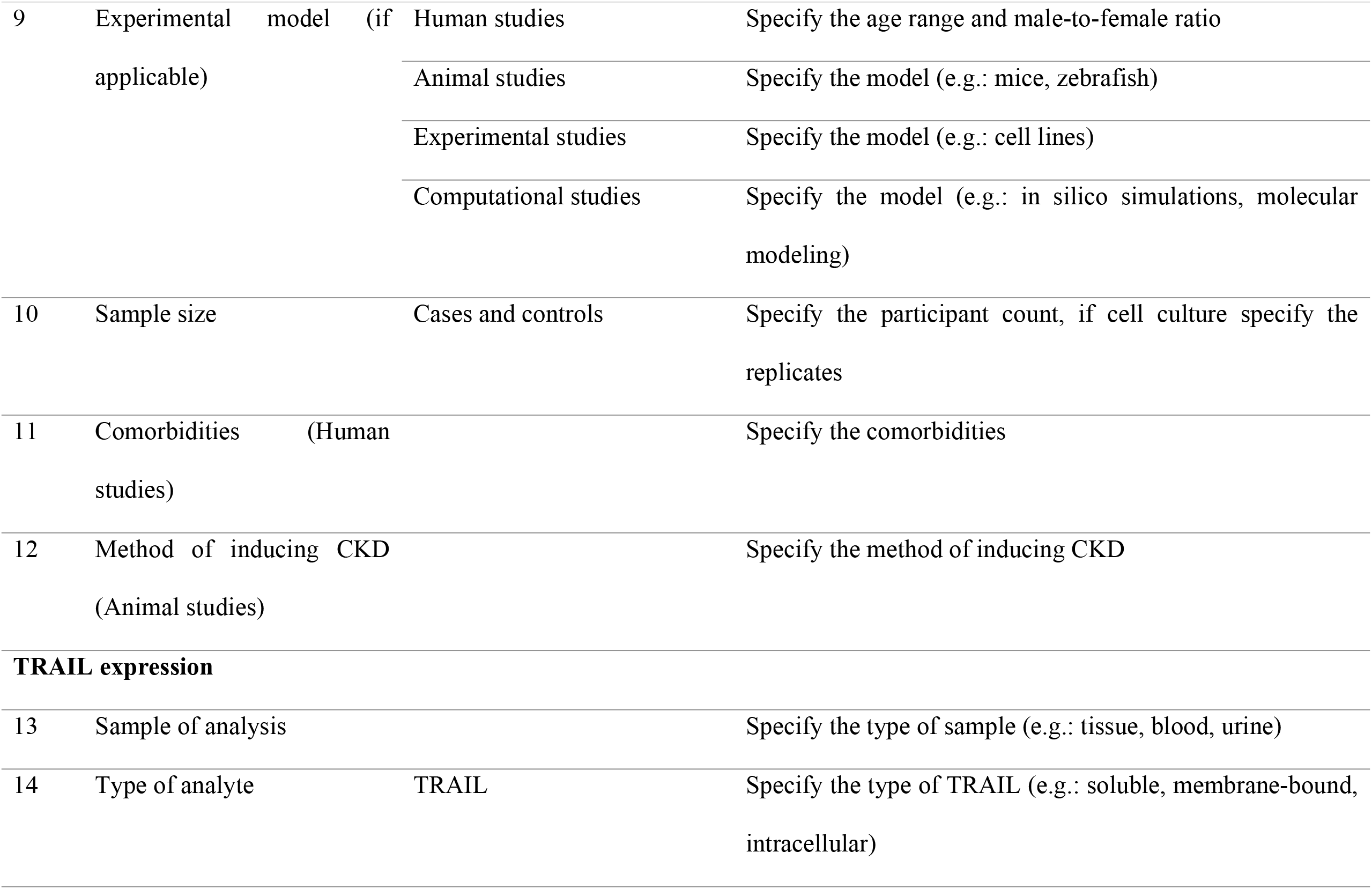
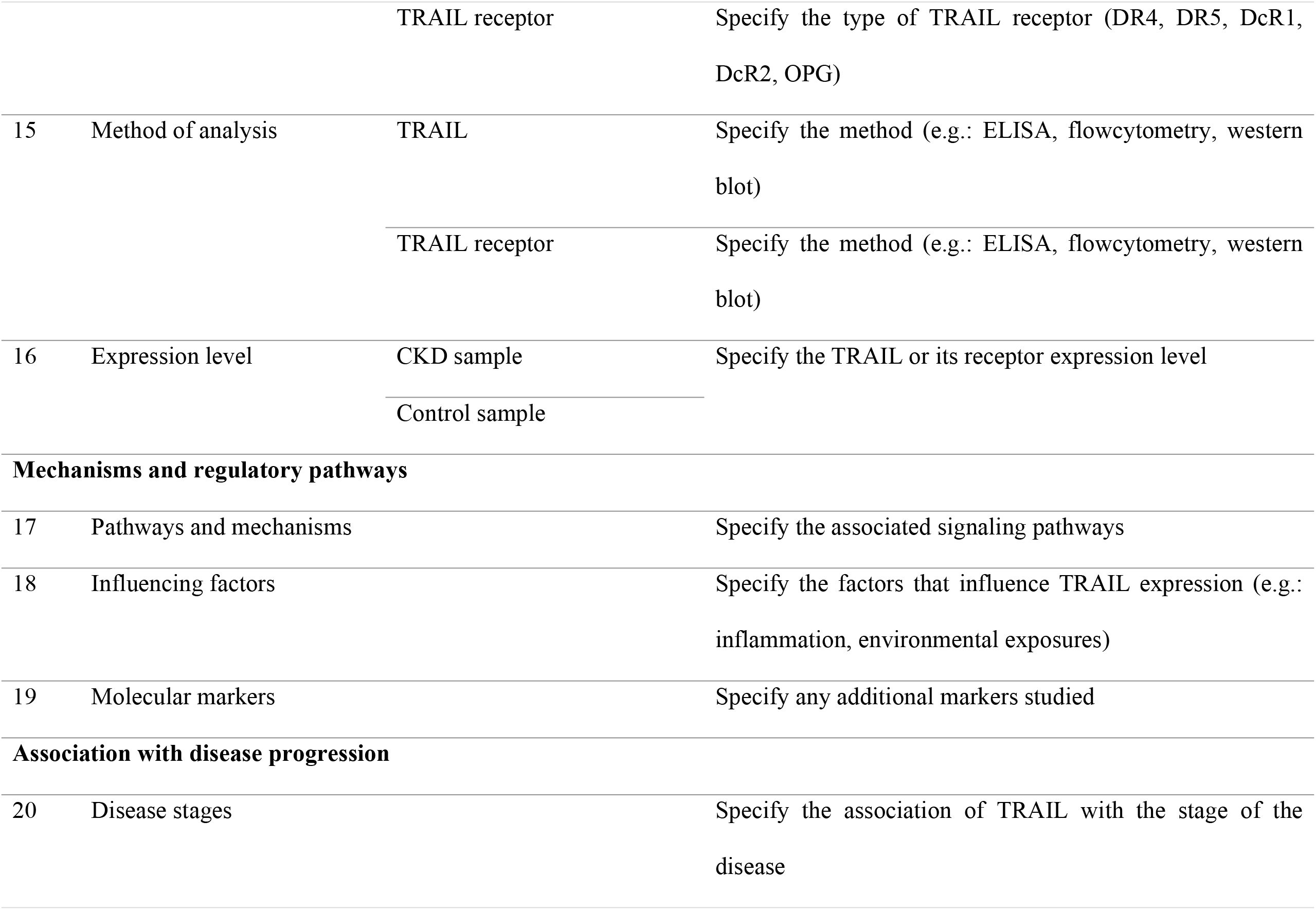
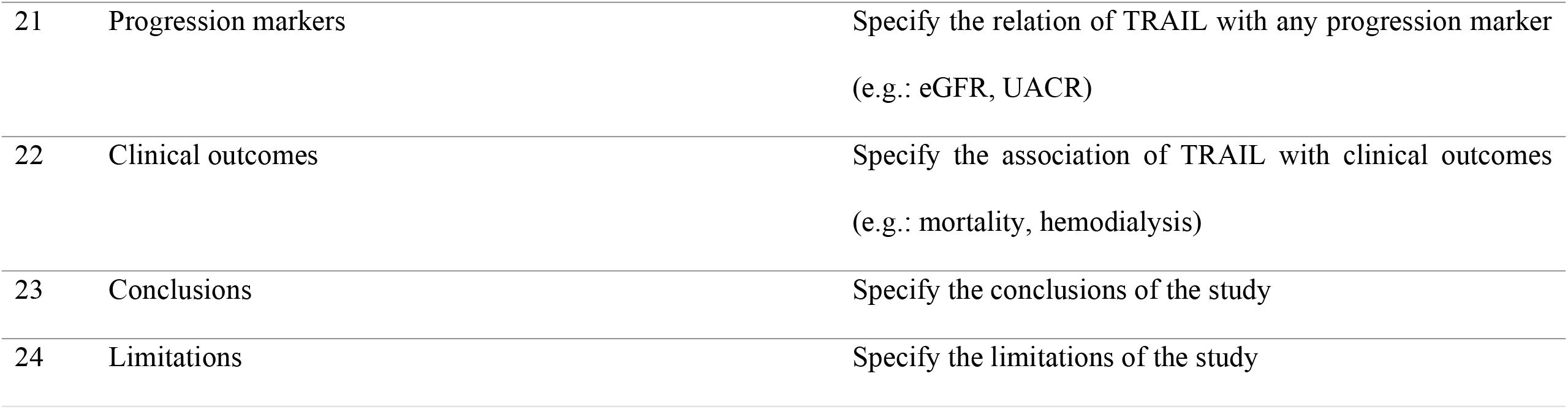
data extraction framework.

1. Study characteristics
2. Population characteristics
3. TRAIL expression
4. Underlying mechanisms and regulatory pathways
5. Association with CKD progression

### Stage 5: Collating and summarizing the results

The outcome of this scoping review will address the research questions mentioned above (section: stage 1). PRISMA flow diagram will be used to illustrate the article selection process including the total number retrieved, duplicates removed, and the final number included. Data analysis will involve tabulating quantitative data and thematically analyzing qualitative data presented narratively or in tables. After contextualizing based on research questions, results of quantitative data analysis will be mainly presented using counts and frequencies, and proportions. The synthesis will be represented through tables, graphs, or charts, complemented by narrative descriptions.

## Data Availability

Data will be made publicly available when the study is completed and published as a scoping review.

## Ethics and dissemination

As this study consolidates secondary data, ethical approval will not be required. The findings will be disseminated in peer-reviewed journals and as presentations at national/international conferences.

## Authors’ contributions

HN contributed to the conceptualization, development of the search strategy, and protocol drafting. HW contributed to the development of the search strategy, drafting, editing, and supervision. NH contributed to the reviewing, editing, and supervision. KW contributed to the conceptualization, development of the search strategy, reviewing and editing the protocol, and supervision. PJ reviewing and supervision.

## Acknowledgement

This work is related to the major project of Science and Technology Human Resource Development Project, Ministry of Higher Education, Sri Lanka, funded by the Asian Development Bank (Grant No. R2RJ4).

## Supporting information

S1 Checklist. Preferred Reporting Items for Systematic Reviews and Meta-Analyses protocol (PRISMA-P) checklist.

